# Experience with Hydroxychloroquine and Azithromycin in the COVID-19 Pandemic: Implications for QT Interval Monitoring

**DOI:** 10.1101/2020.04.22.20075671

**Authors:** Archana Ramireddy, Harpriya Chugh, Kyndaron Reinier, Joseph Ebinger, Eunice Park, Michael Thompson, Eugenio Cingolani, Susan Cheng, Eduardo Marban, Christine M. Albert, Sumeet S. Chugh

## Abstract

**Background:** Despite a paucity of clinical evidence, hydroxychloroquine and azithromycin are being administered widely to patients with verified or suspected COVID-19. Both drugs may increase risk of lethal arrhythmias associated with QT interval prolongation.

**Methods:** We performed a case series of COVID-19 positive/suspected patients admitted between 2/1/2020 and 4/4/2020 who were treated with azithromycin, hydroxychloroquine or a combination. We evaluated baseline and post-medication QT interval (QTc, Bazett’s) using 12-lead ECGs. Critical QTc prolongation was defined as: a) maximum QTc ≥500 ms (if QRS <120 ms) or QTc ≥550 (if QRS ≥120 ms) and b) increased QTc of ≥60 ms. Tisdale score and Elixhauser comorbidity index were calculated.

**Results:** Of 490 COVID-19 positive/suspected patients, 314 (64%) received either/both drugs, and 98 (73 COVID-19 positive, 25 suspected) met study criteria (age 62±17 yrs, 61% male). Azithromycin was prescribed in 28%, hydroxychloroquine in 10%, and both in 62%. Baseline mean QTc was 448±29 ms and increased to 459±36ms (p=0.005) with medications. Significant prolongation was observed only in men (18±43 ms vs -0.2±28 ms in women, p=0.02). 12% of patients reached critical QTc prolongation. In a multivariable logistic regression, age, sex, Tisdale score, Elixhauser score, and baseline QTc were not associated with critical QTc prolongation (p>0.14). Changes in QTc were highest with the combination compared to either drug, with many-fold greater prolongation with the combination vs. azithromycin alone (17±39 vs. 0.5±40 ms, p=0.07). No patients manifested torsades de pointes.

**Conclusions:** Overall, 12% of patients manifested critical QTc interval prolongation, and traditional risk indices failed to flag these patients. With the drug combination, QTc prolongation was several-fold higher compared to azithromycin alone. The balance between uncertain benefit and potential risk when treating COVID-19 patients with these drugs should be carefully assessed prior to use.

## INTRODUCTION

The ongoing COVID-19 pandemic is an unprecedented public health challenge at a global level.^1^ As of April 9, 2020, ≥395,000 Americans have tested positive and over ≥12,700 of those patients^2^ have succumbed to this illness. In the absence of a vaccine or any proven therapeutic agent, hydroxychloroquine (HCQ) and azithromycin (AZ), often used in combination, have emerged as a potential therapy based on extremely limited clinical evidence.^3^ These drugs are currently being prescribed in COVID-19 positive or suspected patients in growing numbers across the US and around the world.^4^

HCQ is an anti-malarial drug which has also been used to treat arthritis and systemic lupus erythematosus. AZ is a macrolide antibiotic used to treat a wide variety of bacterial infections. Both drugs prolong the QT interval by blocking the KCNH2-encoded hERG/Kv11.1 cardiac potassium channel, thereby increasing the risk of torsades des pointes.^5-7^ In general, HCQ is reasonably well-tolerated and used chronically in arthritis and SLE patients without heart rhythm monitoring.^8^ In 2013, the FDA issued a black warning for AZ following multiple reported cases of QTc prolongation followed by torsades des pointes.^9^ Since the use of these drugs in COVID-19 patients is so recent and based largely on *in vitro* studies^10^ and anecdotal observations^11^, community prescribing practices are as-yet unclear. More importantly, there is no information available regarding the extent of QTc prolongation with a combination of HCQ and AZ. From first principles, simultaneous block of the hERG/Kv11.1 cardiac potassium channel with both agents may cause critical QTc prolongation and an elevated risk of torsades des pointes. We therefore evaluated prescribing practices and monitored the QTc interval in COVID-19 positive or suspected patients who received these medications.

## METHODS

### Patient Population

This study was approved by the Cedars-Sinai Institutional Review Board. We identified all patients with confirmed COVID-19 infection as well as patients under investigation (PUI) admitted at Cedars-Sinai Medical Center in Los Angeles, California between 2/1/2020 and 4/4/2020 who received either AZ and/or HCQ as part of their medical treatment. A detailed, retrospective chart review was then performed via the electronic health record (Epic, Verona, WI) at our institution. Of the PUI and confirmed COVID-19 cases, we only included patients who had at least two 12-lead electrocardiograms (ECGs) performed in our MUSE system between 1/1/2020 and 4/5/2020. Patients with paced ventricular rhythms, atrial fibrillation, atrial flutter, supraventricular tachycardia, or ECGs otherwise unsuitable for accurate QT interval measurement were excluded.

Additionally, patients without ECGs performed on day 2 of medication administration or later were excluded (Figure 1). Our hospital policy states that daily ECGs should be performed in all patients who are started on the combination of AZ and HCQ just prior to and for the duration of therapy. If a patient has QTc ≥ 470 ms at baseline or reaches critical QTc prolongation (defined below in Outcomes), then continuous telemetry monitoring is recommended.

**Figure 1.**
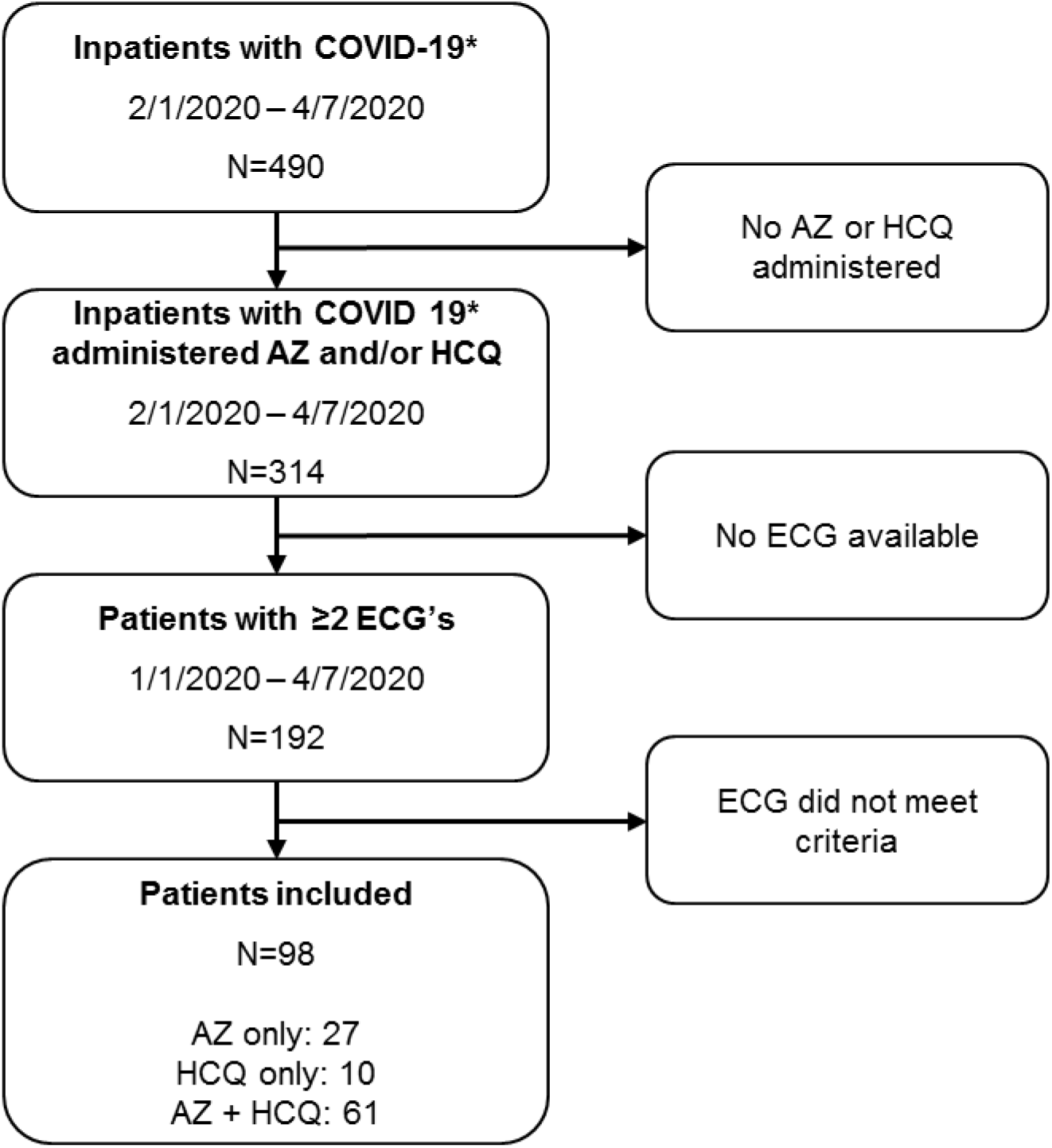
Study Design. *COVID-19 positive patients and patients under investigation (PUI). AZ=Azithromycin, HCQ=Hydroxychloroquine.

### Data Collection and Definitions

We compared obtained standard 12-lead ECG parameters, including the corrected QT interval (QTc, Bazett’s formula) from the baseline ECG closest to and prior to medication administration, to the longest QTc observed post-medication administration up to 24 hours after the last medication dose. Given variability in heart rates, QTc prolongation was also evaluated using the Fridericia formula for correction of the QT interval. QT measurements were independently reviewed and validated by a cardiologist to ensure accuracy of the automatic measurement and manually re-measured if needed. Patients were assigned into to one of 3 three groups depending on which medication they received – AZ, HCQ, or AZ + HCQ. Patient demographics, clinical history, labs, and medications were obtained from detailed chart review, including risk factors to calculate the Tisdale score^12^ and the Elixhauser comorbidity index^13^ for each patient. The Tisdale score consists of age ≥ 68 yrs, female sex, use of loop diuretics, potassium level ≤ 3.5 mEq/L, baseline QTc ≥ 450 ms, acute MI, number of QT prolonging medications, sepsis, and heart failure and has been developed as a risk score for QTc interval prolongation. The Elixhauser comorbidity index incorporates 21 different variables, including obesity, diabetes, peripheral artery disease, chronic kidney disease, HIV, and cancer and is used to predict in-hospital mortality.

### Outcome

Outcomes of interest were baseline QTc ≥470 ms^14^, and post-medication critical QTc prolongation defined as follows: a) maximum post-medication QTc ≥500 ms (if QRS <120 ms) or QTc ≥550 (if QRS ≥120 ms) and b) mean increase from baseline QTc to maximum post-medication QTc of ≥60 ms.^14, 15^

### Statistical Analysis

We evaluated the mean change from baseline QTc to longest QTc overall and by medication type using repeated measures analysis of variance. We also evaluated the frequency of critical ECG changes overall, by medication type, and by patient characteristics using chi-square analyses. Given the relatively low number of patients who received HCQ alone, the decision was made to only compare AZ to the combination of AZ + HCQ. We used multivariable logistic regression to evaluate whether age, sex, Tisdale score, Elixhauser score, and baseline QTc were associated with critical QTc prolongation.

## RESULTS

A total of 490 patients were admitted to our medical center with a verified or suspected diagnosis of COVID-19 during the study time period, and 314 (64%) were treated with AZ, HCQ, or the combination. Of these, 192 patients had a 12-lead ECG performed during treatment, and a subgroup of 98 confirmed COVID infection or PUI (age 62±17 yrs, 61% male) met criteria for inclusion in the study (Table 1). Ninety-two of the excluded patients either had ECGs with indeterminate QT intervals (see Methods) or no ECG on at day 2 of medication administration or later, and two patients had extreme variation in heart rate, with near doubling of heart rate between baseline and maximal QTc prolongation. Given the limitations of QT interval correction at significantly elevated heart rates, we excluded both patients from the final analysis. Only 1 patient included in the analysis required manual QT measurement using the MUSE electronic calipers and the tangent method of calculation. Of the 98 patients, 73 patients were COVID-19 positive, and 25 were PUI. AZ was prescribed in 28%, HCQ in 10%, and the combination in 62%.

**Table 1.** Patient Characteristics.

| **Characteristics** | **Confirmed COVID/PUI (n=98)** |
| --- | --- |
| Mean age (yr) | 62.3±17.0 |
| Male sex (%) | 60 (61%) |
| Body mass index (BMI) | 27.8±6.6 |
| Ethnicity |  |
| Hispanic | 15 (15%) |
| Non-Hispanic | 79 (81%) |
| Unknown | 4 (4%) |
| Race |  |
| Asian | 5 (5%) |
| Black/African American | 17 (17%) |
| White | 69 (70%) |
| Other | 5 (5%) |
| Unknown | 2 (2%) |
| Heart Failure | 20 (20%) |
| Hypertension | 59 (60%) |
| Diabetes Mellitus | 22 (22%) |
| Chronic Kidney Disease | 14 (14%) |
| Chronic Obstructive Pulmonary Disease (COPD) | 25 (26%) |
| Number of ICU Patients | 48 (49%) |
| Number of non-ICU Patients | 50 (51%) |
| Medications |  |
| Azithromycin (AZ) | 27 (28%) |
| Hydroxychloroquine (HCQ) | 10 (10%) |
| Azithromycin + Hydroxychloroquine (AZ + HCQ) | 61 (62%) |
| Tisdale Score |  |
| Low Risk (<7) | 1 (1%) |
| Moderate Risk (7-10) | 36 (37%) |
| High Risk (≥11) | 61 (62%) |
| Elixhauser Comorbidity Index | 15.2±13.2 |

At baseline, the mean QTc was 448±29ms, and 20% of patients had QTc >470ms (Table 2). With drug administration, overall QTc increased to 459±36ms (p=0.005). A similar increase in QTc was observed using correction of the QT interval with the Fridericia formula (420±32 ms to 438±36, p<0.0001). Subgroup analysis by sex revealed baseline and post-drug QTc of 449.1±31.4ms and 467.3±37.8ms in men (p=0.002). In women, baseline and post-drug QTc were 444.9±24.1ms and 444.7±28.6ms (p=0.97). Significant prolongation was observed only in men versus women (18±43 ms vs -0.2±28 ms, p=0.02). Further analysis by sex showed no significant difference in Tisdale score or Elixhauser comorbidity index. High Tisdale scores were similar in men and women (62% vs 63%, p=0.88). The average Elixhauser comorbidity index in men was lower and non-significant when compared with women. A trend towards significance was noted when comparing overall BMI in men vs women – 42.4% of men were obese vs 26.3% of women (p=0.11).

**Table 2.** ECG Characteristics.

| **Characteristics** | **Confirmed COVID/PUI (n=98)** |
| --- | --- |
| Baseline ECG |  |
| Ventricular Rate (bpm) | 92±19 |
| RR (ms) | 681±148 |
| QRS (ms) | 92±19 |
| QT (ms) | 368±44 |
| QTc (ms, Bazett) | 448±29 |
| Post-Drug ECG |  |
| Ventricular Rate (bpm) | 82±19 |
| RR (ms) | 768±180 |
| QRS (ms) | 94±19 |
| QT (ms) | 400±48 |
| QTc (ms, Bazett) | 459±36 |
| Mean Change in QTc (ms) | 11.1±38.5 |
| Pts with Baseline QTc ≥470ms | 20 (20%) |
| Pts Meeting Critical QTc Threshold |  |
| ≥500ms (QRS <120ms) | 7 (7%) |
| ≥550ms (QRS ≥120ms) | 1 (1%) |
| Pts with Critical Absolute Change in QTc |  |
| ΔQTc ≥60ms and QTc <500ms/550ms threshold | 4 (4%) |
| ΔQTc ≥60ms and QTC ≥500ms/550ms threshold | 7 (7%) |
| Total Pts with Critical QTc Prolongation | 12 (12%) |

Of note, multiple patterns were observed for use, duration, and dosage of both medications. Though a majority (81.6%) of patients received at least a 5-day course of each medication, there were many patients who received a higher dose on day 1 compared to the dosage on days 2-5. For example, AZ was either given as PO or IV, and the dosage was either 500mg daily or 500mg on day 1 followed by 250mg daily on days 2-5. For HCQ, most patients (87.0%) received 400mg PO twice daily prior to receiving a dosage of 200mg PO twice daily on days 2-5. It was noted that the medications were either started together (50.8%), staggered by one day (29.5%), or administered more than one day after the other (19.7%). Patients with lower values of QTc at baseline were more likely to be started on the combination of the two medications. In contrast, patients with a higher baseline QTc were more likely to receive either HCQ or AZ rather than the combination.

The mean change in QTc values was greatest in the combined HCQ and AZ group, and when compared to the AZ group, QTc prolongation due to the combination was observed to be several-fold higher than due to AZ alone (17.2±39.0 ms vs 0.5±40.3 ms, p=0.07) (Figure 2). A subgroup of 12% of patients were observed to reach a critical level of QTc prolongation – either QTc ≥ 500ms (if QRS < 120ms), QTc ≥ 550ms (if QRS ≥ 120ms), or change in QTc ≥ 60ms (Figure 3). Among the 12 patients with critical QTc prolongation, 5 patients were on AZ, and 7 patients were on the combination of HCQ and AZ (Table 3).There were no significant differences in regards to age, sex, sepsis, diabetes, baseline QTc ≥450 ms, or number of QT prolonging medications. Patients with critical QTc prolongation had significantly greater use of loop diuretics (p=0.007) and acute MI (p=0.01) (Table 4). 75% of patients with critical QTc prolongation had a high Tisdale score whereas 60.5% of patients without critical QTc changes had a high Tisdale score (p=0.33). Similarly, there was no significant difference in the Elixhauser comorbidity indices of the two groups (p=0.86). In a multivariable logistic regression, none of the following predictors were significantly associated with critical QTc prolongation: age, sex, Tisdale score, Elixhauser score, and baseline QTc (p>0.14). No patients had syncope, torsades de pointes, or other lethal arrhythmias during or after drug administration.

**Table 3.** Baseline and Post-Drug ECG Characteristics by Medication Administered.

| ECG Characteristics | Azithromycin<br>(n=27) | Azithromycin +<br>Hydroxychloroquine<br>(n=61) | P<br>value |
| --- | --- | --- | --- |
| Baseline ECG |  |  |  |
| Vent Rate (bpm) | 102±23 | 89±17 | **0.01** |
| RR (ms) | 622±168 | 701±135 | **0.02** |
| QRS (ms) | 94±24 | 93±17 | 0.84 |
| QT (ms) | 364±61 | 367±37 | 0.84 |
| QTc (ms, Bazett) | 463±39 | 439±20 | **0.005** |
| Post-Drug ECG |  |  |  |
| Vent Rate (bpm) | 92±23 | 78±15 | **0.006** |
| RR (ms) | 686±168 | 794±148 | **0.003** |
| QRS (ms) | 93±24 | 94±18 | 0.76 |
| QT (ms) | 383±62 | 405±43 | **0.10** |
| QTc (ms, Bazett) | 464±38 | 457±38 | 0.41 |
| Change in QTc (ms) | 0.5±40.3 | 17.2±39.0 | **0.07** |
| Pts Meeting Critical QTc Threshold* | 3 (11%) | 5 (8%) | 0.66 |
| Pts with Absolute ΔQTc ≥60ms | 4 (15%) | 7 (12%) | 0.66 |
| Pts with Critical QTc Prolongation Overall† | 5 (19%) | 7 (12%) | 0.37 |

\* ≥ 500ms (QRS <120ms) or ≥ 550ms (QRS ≥ 120ms)
† By QTc threshold or absolute change criteria

**Table 4.** Characteristics of Patients with Critical QTc Prolongation.

|  | **Critical QTc**<br>**Prolongation**<br>**(n=12)** | **Non-Critical**<br>**Change in QTc**<br>**(n=86)** | **P**<br>**value** |
| --- | --- | --- | --- |
| **Tisdale Score** |  |  |  |
| Age ≥ 68 yrs | 3 (25%) | 39 (45%) | 0.18 |
| Female Sex | 3 (25%) | 35 (41%) | 0.29 |
| Loop Diuretic | 6 (50%) | 14 (16%) | **0.007** |
| Serum K<sup>+</sup> ≤ 3.5 mEq/L | 1 (8%) | 18 (21%) | 0.30 |
| Baseline QTc ≥ 450 ms | 3 (25%) | 38 (44%) | 0.21 |
| Acute MI | 5 (42%) | 11 (13%) | **0.01** |
| ≥2 QT Prolongs Drugs | 11 (92%) | 63 (73%) | 0.16 |
| Sepsis | 9 (75%) | 62 (72%) | 0.83 |
| Heart Failure | 3 (25%) | 17 (20%) | 0.67 |
| **Elixhauser Index** |  |  |  |
| COPD* | 2 (17%) | 23 (27%) | 0.45 |
| Diabetes Mellitus | 3 (25%) | 19 (22%) | 0.82 |
| Hypertension | 7 (58%) | 52 (61%) | 0.89 |
| CKD<sup>†</sup> | 3 (25%) | 11 (13%) | 0.26 |
| **Male Sex** | 9 (75%) | 51 (59%) | 0.29 |
| **Low/Moderate Tisdale Score** | 3 (25%) | 34 (40%) | 0.33 |
| **High Tisdale Score** | 9 (75%) | 52 (61%) | 0.33 |
| **Elixhauser Index** | 14.6±9.2 | 15.3±13.7 | 0.86 |

\* COPD = chronic obstructive pulmonary disease
<sup>†</sup> CKD = chronic kidney disease

**Figure 2.**
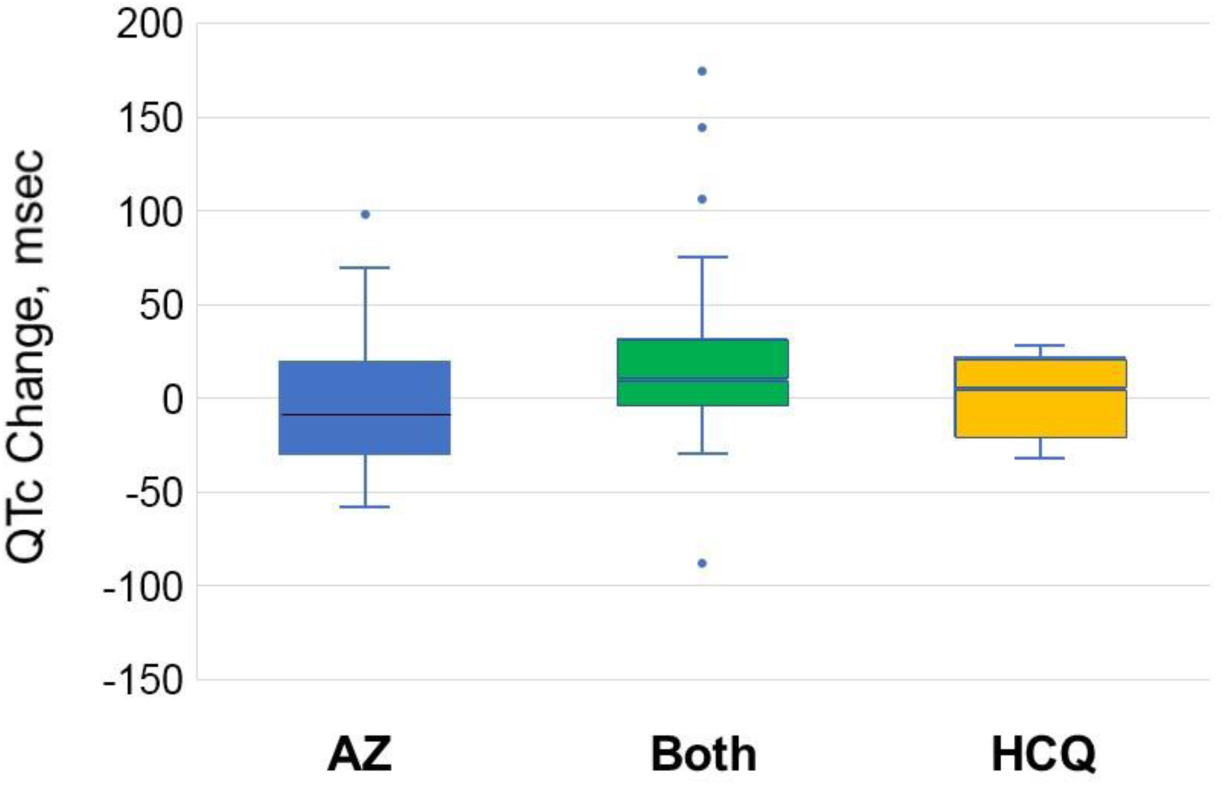
Boxplot of QTc changes by medication group. The QTc change is greatest in the combined HCQ and AZ group, and when compared to the Azithromycin group alone, the change in QTc is borderline significant (p=0.07). AZ=Azithromycin, HCQ=Hydroxychloroquine.

**Figure 3.**
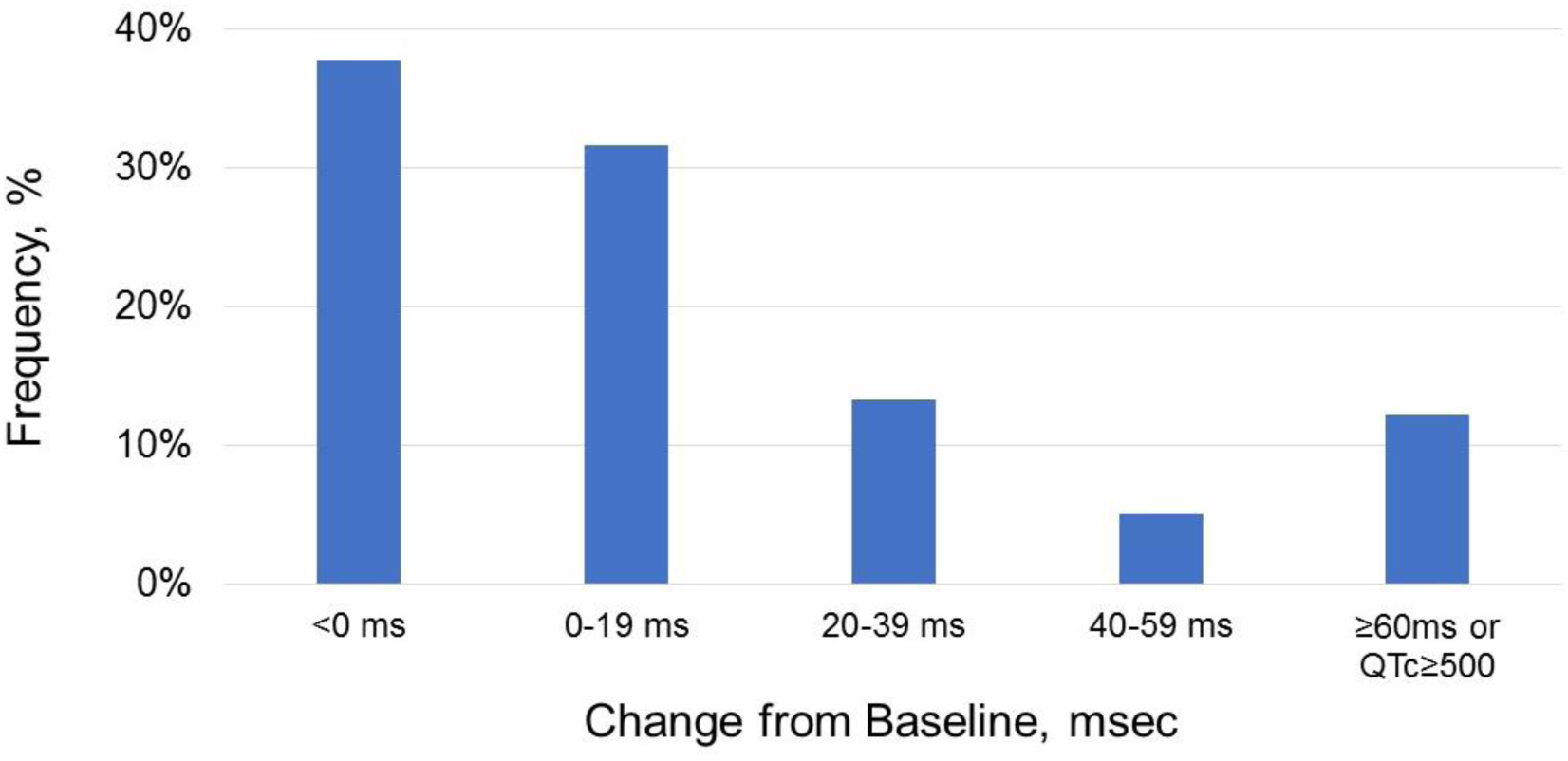
Change in QTc from baseline to post-medication administration. Critical QTc prolongation (Δ ≥60ms or QTc ≥500ms (≥550ms in pts with QRS≥120ms)) was observed in 12% of patients.

## DISCUSSION

At a single large community medical center, we studied 98 COVID-19 positive or suspected patients with HCQ and AZ, administered either individually or in combination. Although lethal arrhythmias were not observed during the limited period of observation, 12% of patients attained critical QTc prolongation, and that QTc prolongation was many-fold higher when HCQ and AZ were administered in combination compared with AZ alone. Moreover, patients who achieved critical QTc prolongation were not distinguished by significantly higher Tisdale scores, nor by higher Elixhauser comorbidity indices.

Our finding of greatest QTc prolongation in the combined HCQ + AZ group is important. In our study, patients with lower values of QTc at baseline were more likely to be started on the combination of the two medications. In contrast, patients with a higher baseline QTc were more likely to receive either HCQ or AZ rather than the combination. This suggests that prescription practice may have been driven by a concern regarding excessive QTc prolongation with the use of both medications in combination. Further review of our data showed that some practitioners decided on a staggered dosing regimen of one drug followed by the other drug later in the hospital stay. However, given the prolonged half-lives of these medications, such practice still results in a combined drug effect on the QTc.

Another interesting finding in our study was that men had significantly higher rates of QTc prolongation than women. The incidence of torsade de pointes is known to be higher in women, and female sex is a risk factor in the Tisdale score for QT prolongation. The reasons for this discrepancy still need to be explained, but could be related to the majority of patients in our population being men. Since rates of high Tisdale scores and the Elixhauser comorbidity index were also not significantly varied between the sexes, this finding cannot be explained by differences in overall morbidity burden and would benefit from further study.

Overall, our findings support the need to monitor the QTc during the period that these two medications are administered, especially when used in combination. We were not able to distinguish patients more likely to have critical QTc prolongation by the Tisdale score or Elixhauser index, making monitoring during medication administration likely to be a safer approach. ^14^ In addition to QTc prolongation as a result of KCNH2-encoded hERG/Kv11.1 cardiac potassium channel blockage, both drugs can provoke potentially lethal arrhythmias by additional mechanisms.^16, 17^ In fact, polymorphic ventricular tachycardia with AZ has been reported in the absence of QTc prolongation.^18^ Given that critical QTc prolongation was observed with both administration of AZ (n=5) as well as the combination of HCQ + AZ (n=7), monitoring of the QTc interval should not be restricted to the combination of these drugs alone. This is consistent with the 2013 black box warning issued by the FDA regarding the increased risk of torsades de pointes with AZ.

### Limitations

The variation in dosing patterns and duration of treatment for each medication are important to note as this can potentially influence when the ECG with the longest QTc may be observed. Due to inconsistency in obtaining daily ECGs during medication administration, we believe that the 12% of patients who experienced critical QTc prolongation was likely an under-estimation. Furthermore, variability in heart rates, especially in the setting of sepsis, can affect QT interval correction. We reported the QTc using Bazett’s correction, which is consistent with the majority of the published literature as well as ECG recording machines in typical use. The relatively small sample size does not allow for assessment of torsades des pointes risk with these agents.

## CONCLUSION

Prescribing practices for AZ and HCQ amid the COVID-19 pandemic were found to be diverse, but the majority of patients in our study received a combination of the two drugs. Among patients prescribed AZ, HCQ, or a combination of both, 12% of patients achieved a critical level of QTc interval prolongation. QTc values were highest with the combination of HCQ and AZ, with QTc prolongation many-fold higher in the combination group than with AZ alone. Especially if ongoing clinical trials indicate benefit from use of these medications, these findings could inform clinical therapeutics and ECG monitoring in COVID-19 patients.

## Data Availability

No external datasets or supplementary material online was utilized. All data originated from Cedars-Sinai Medical Center.

